# Socioeconomic inequalities in blood pressure: co-ordinated analysis of 147,775 participants from repeated birth cohort and cross-sectional datasets, 1989 to 2016

**DOI:** 10.1101/2019.12.19.19015313

**Authors:** David Bann, Meg Fluharty, Rebecca Hardy, Shaun Scholes

## Abstract

**Objective:** To evaluate whether socioeconomic inequalities in blood pressure (BP) have changed from 1989 to 2016—a period in which average BP levels declined and the detection and treatment of high BP increased.

**Design:** Repeated observational studies.

**Setting:** 3 nationally representative British birth cohort studies—born in 1946, 1958, and 1970—and 21 nationally representative cross-sectional studies (Health Survey for England 1994 to 2016, HSE).

**Participants:** 147,775 participants with BP outcomes at age 42-46 years (cohorts) or 25 years and over (HSE).

**Main outcome measure:** Absolute differences in systolic BP (SBP) by educational attainment (cohorts and HSE) and early life social class (cohorts).

**Results:** In both datasets, lower education was associated with higher SBP, with similar absolute magnitudes of inequality across the studied period. Differences in SBP by education (Slope Index of Inequality) based on HSE data were 3.0mmHg (95% CI: 1.8, 4.2) in 1994 and 4.3mmHg (2.3, 6.3) in 2016. Findings were similar for diastolic BP (DBP) and survey-defined hypertension. Cohort data suggested that disadvantage in early and adult life had cumulative independent associations with BP: cohort-pooled differences in SBP were 4.9mmHg (3.7, 6.1) in a score combining early life social class and own education, yet were 3.4mmHg (2.4, 4.4) for education alone. In both datasets, inequalities were found across the SBP distribution—below and above hypertension thresholds—yet were larger at the upper tail; in HSE, median SBP differences were 2.8mmHg (1.7, 3.9) yet 5.6mmHg (4.9, 6.4) at the 90th quantile.

**Conclusion:** Socioeconomic inequalities in BP have persisted from 1989 to 2016, despite improved detection and treatment of high BP. To achieve future reductions in BP inequalities, policies addressing the wider structural determinants of high BP levels are likely required—targeting detection and treatment alone is unlikely to be sufficient.

## Introduction

Blood pressure (BP) is a key modifiable determinant of cardiovascular disease, with higher risk continuing across its distribution beyond the thresholds for initiating treatment.^1^ Cardiovascular diseases in turn remain leading causes of mortality worldwide,^2^ with substantial public health and associated economic costs. Higher BP—particularly in midlife^3 4^—is also implicated in the occurrence of dementia and related neurological outcomes.^5 6^ As such, monitoring of BP at the population-level is important, as are understanding its socioeconomic inequality given its likely contribution to inequities in cardiovascular disease morbidity and mortality.^7^

While there is repeated evidence that more disadvantaged socioeconomic position (SEP) is correlated with, and potentially causally linked with, higher BP,^8-13^ important gaps in understanding remain. Effects of SEP on a given health outcome are likely to be context-specific and modifiable by societal change and public health policy.^14 15^ In the UK and other high income countries, average BP levels have declined from the 1970s onwards^16^—a change typically attributed to decreased salt intake in foods^17^ and improved hypertension detection and treatment.^18^ How these and other changes such as secular increases in diabetes and obesity (themselves socially patterned)^12 19^ have impacted on socioeconomic inequalities in BP however, remains unclear. The most recent UK analysis of trends in BP inequalities used repeated cross-sectional data from England in 1994-2008^12^ and requires updating using more recent data. It is also likely limited by 1) using an indirect area-based SEP indicator, which may be biased due to measurement error or socioeconomic and health-related migration,^20 21^ and 2) not accounting for use of BP lowering treatment—an important consideration since increases in levels of treatment for high BP across time in England and worldwide^18 22^ may have narrowed or widened observed (post-treatment) inequalities in BP.

There are also uncertainties regarding the life course nature of inequalities in BP—that is, when in life, exposure to disadvantaged SEP impacts on adult BP outcomes. Existing studies have suggested that there are cumulative effects of disadvantaged SEP across life on higher BP (i.e. that there are independent effects of exposure to disadvantage in both child and adult life),^23^ while other studies have suggested that childhood SEP is particularly important for adult BP (among men).^24^ This evidence is based on analysis conducted in separate British birth cohorts using different empirical strategies.^23-26^ In contrast, a co-ordinated analytic approach across these cohorts can increase statistical power, improve result generalisability, and enable testing of whether such processes have changed across time.

A final limitation of the existing evidence—across all data sources—is that studies typically examine SEP differences in average levels of BP (linear regression), or in binary outcomes (logistic regression);^13^ it is therefore unclear how SEP differences relate to the population distribution of BP. The consequences of BP inequalities are likely more pronounced if the magnitude of associations are largest amongst those at highest risk—i.e., those with highest BP levels (the upper tail of the distribution). While infrequently used in the epidemiological literature, quantile regression analysis can inform this.^27^

UK studies of BP inequalities have used different data sources separately—such as repeated cross-sectional^12^ or single birth cohort studies.^10 24 25 28 29^ This study is the first, to our knowledge, to conduct co-ordinated analyses across national birth cohort studies and repeated cross-sectional surveys to examine change across time in BP inequalities. A co-ordinated approach across these different data sources has multiple advantages. First, given their independent sampling designs, the robustness of the results and generalisability to the population is likely improved. Second, they have complementary strengths which aid understanding—birth cohort studies contain prospective SEP data across life, yielding insight into when in life exposure to disadvantage may affect BP outcomes in adulthood. The availability of BP measures in midlife in each birth cohort (age 42-46 years) enables sufficiently powered investigation of how inequalities have changed across time at this key life stage.^3 4 30^ Whilst lacking information on early life SEP, use of nationally representative repeated cross-sectional data enable investigation of how inequalities in BP have changed across all adults and period (annually from 1994). Consistent with a fundamental cause model of health inequality,^31^ we hypothesised that inequalities in BP have persisted across the studied period (1989 to 2016) since there are multiple likely mediating pathways between SEP and BP, some of which are likely to have stubbornly persisted over the last two decades (e.g., diabetes and obesity^12 19^). We also hypothesised that exposure to disadvantage in both early and adult life would have persisting additive cumulative associations with higher BP. Finally, we hypothesised that such inequalities would be largest at the upper tail of the BP distributions, as previously found in the UK and elsewhere for body mass index (BMI).^27^

## Methods

### Study samples

For each dataset, participants gave verbal and/or written consent to be interviewed, visited by a nurse, and to have BP measurements taken during a home visit. Research ethics approval was obtained from relevant committees.

### Birth cohorts

Britain’s birth cohort studies—followed-up to adulthood—were used. These were designed to be nationally representative when initiated in 1946 (MRC National Survey of Health and Development^32 33^—1946c), 1958 (National Child Development Study^34^—1958c), and 1970 (British Cohort Study^35^— 1970c). The history, design, and characteristics of these studies have been previously described in detail in papers^32-36^ and books;^37 38^ studies have also examined the characteristics of those lost due to attrition.^39-42^ To aid the comparability of associations between SEP and BP across studies, analyses were restricted to singleton births in mainland Britain from those born and included in cohorts in the relevant weeks in March/April 1946c (N=5362), 1958c (N=16 989), and 1970c (N=16 216). Subsequent migrants included in childhood sweeps of 1958c and 1970c were not included. The analytic sample sizes were those with valid BP data from age 42–46 years—total N=18 657. Cohort-specific analytic sample sizes (Ns for those who responded but did not have valid BP data are in parenthesis) are as follows: 1946c: n=3186 (3262); 1958c: n=8610 (8688); 1970c: n=6861 (7793). In 1970c, unlike the other cohorts, nurse visits took place following separate main stage interviews. Age ranges at BP measurement in midlife were: 1946c: 42–44 (1988-1990; <1% in 1988, thus 1989 is referred to throughout as the start date of analyses); 1958c: 44–46 (2002-2004); 1970c: 45–48 years (2015-2018). All analyses using the 1946c were weighted to account for the stratified sampling design.^33^ To reduce the potential impact of missing data on statistical power and the potential for selection bias in longitudinal analyses, missing SEP data for those with valid BP data were replaced using multiple imputation (n=10 datasets; analyses using imputed data are presented).^43^

### Repeated cross-sectional studies

The Health Survey for England (HSE) is a cross-sectional, general population survey of individuals living in private households, with a new sample each year randomly selected by address.^44^ Data collection occurs throughout the year, via a health interview followed by a nurse visit. Twenty-one sweeps of the HSE were used for the present analysis spanning 1994 to 2016 (no BP measurements for participants in the general population sample were available in 1999 or 2004). The percentage of eligible households taking part in the HSE ranged from 77% in 1994 to 59% in 2016. The analytical sample included n=129 118 non-pregnant adults aged 25 years and over with valid BP, medication and education data; 25 was chosen as the lower age limit to avoid bias in the classification of highest educational attainment (see below). This represented 65% of all adult participants. Our study used the general population samples only and so selection probabilities were equal. Starting from 2003, weights have been created to minimise bias from non-response; the relevant weights for analysing BP data were therefore used from 2003 onwards. Sensitivity analyses suggested no systematic difference in results for weighted versus unweighted data.

### Measurement of blood pressure

#### Birth cohorts

Trained nurses measured resting BP using a sphygmomanometer following a rest time of 5 minutes. Specifically, in 1946c BP was measured twice using the Hawksley random zero sphygmomanometer in the right arm; in 1958c three times using an Omron 705CP BP monitor in the left arm; and in 1970c three times using an Omron HEM 907 BP monitor in the right arm. To account for differences in device, as Omron devices provide higher estimates than sphygmomanometers, the 1946c readings were converted to automated Omron readings via a previously derived formulae.^45^ Pregnant women were excluded from BP measurement in the 1958c and 1970c. To aid within-cohort comparability, the second BP measure was used, or the first if the second was missing. Use of medication was ascertained in each cohort which was categorized as either BP lowering (1) or not (0); this was derived using a single item on self-reported use of hypertension medication in 1946c and coded from medication data in 1958c and 1970c using British National Formulary codes.^46^

### Repeated cross-sectional studies

BP was measured with the use of Dinamap 8100 monitors before 2003, and Omron HEM 907 from 2003 onwards.^22^ We converted Dinamap readings into Omron readings using a regression equation based on a calibration study.^22^ Across all survey years, three BP readings were taken from each participant in a seated position at 1 minute intervals with use of an appropriately sized cuff on the right arm if possible after a 5 minute rest. Participants who had exercised, eaten, drunk alcohol, or smoked in the 30 minutes before measurements were excluded from analyses. The mean of the second and third readings was used across all survey years. Details of which, if any, classes of antihypertensive medicines were being taken were recorded by the nurse.

### Socioeconomic position ascertainment

Given evidence in a recent meta-analysis for the particular relevance of education for social gradients in BP,^13^ and evidence for its potential causal role,^7 9^ it was the main SEP indicator in this study. Education for HSE participants was measured as the highest qualification attained (at the time of interview). For the birth cohort studies, own highest education attainment was ascertained at ages 26 (1946c), 34 (1958c), and 33 (1970c). For both data sources, highest educational attainment at the time of measurement was categorised into: degree/higher, A levels/diploma, O Levels/GCSEs/Vocational equivalent, or none. In addition, a comparable indicator of early life (childhood) SEP was used in analyses of birth cohort data. This was measured by father’s social class at age 4 in 1946c (occupation at birth was not used to avoid WWII-related misclassification), and at birth in 1958c and 1970c. Data at 10/11 years was used if missing at the earlier age. The Registrar General’s Social Class scale was used with categories as follows— I (professional), II (managerial and technical), IIIN (skilled non-manual), IIIM (skilled manual), IV (partly-skilled), and V (unskilled) occupations. To examine cumulative associations (see below), the ridit scores for social class at childhood and own education were combined into a single score and re-scaled.

### Analytical strategy

The analytical sample size was 147 775—those with BP outcomes at age 42-46 years (cohorts) or 25 years and over (HSE). The primary outcome for our study was the continuous level of systolic BP (SBP). In addition, analyses were repeated for diastolic BP (DBP) and the binary outcome of survey-defined hypertension (SBP≥140mmHg or DBP≥90mmHg or reported use of BP lowering medication). A constant of 10mmHg (SBP) and 5mmHg (DBP) was added to the observed BP values amongst those on BP lowering medication to approximate underlying BP (i.e. the BP individuals would have if they were not on treatment);^47^ this method has been found to reduce bias in the estimated effect of key determinants on continuous levels of BP due to the effects of treatment.^48^

The birth cohort and HSE datasets were analysed separately. Data from each source was analysed overall (i.e., pooled across cohorts / survey years) as well as separately by birth cohort / survey year. To provide single quantifications of inequalities in a metric recommended for use in national health inequality statistics,^49^ the SEP indicators described above were converted to ridit scores (ranging from 0 to 1). The SEP coefficient in linear regression—the Slope Index of Inequality (SII)—is interpreted as the estimated absolute (mean) difference (absolute inequality) in BP between the lowest and highest SEP. For survey-defined hypertension, this forms a linear probability model^50^—the absolute difference in the probability of hypertension between the lowest and highest SEP; our findings did not differ materially when estimated using logistic regression. Use of ridit scores enables comparisons across birth cohorts and survey years whilst accounting for differences in the proportion of participants in each SEP category. Given the large age range in the HSE datasets, adjustment was made for age in all models (entered as a categorical variable via 10-year age bands). No adjustment was made in the birth cohort datasets as the age differences were small and were co-linear with birth year. Our regression models were also gender-adjusted, since gender differences in SEP and BP associations were not anticipated a-priori. Investigation of change across time in the SEP and BP associations was performed by comparing the magnitude (and 95% Confidence Intervals) of the birth cohort and survey-year specific estimates.

### The life course nature of inequalities in BP: birth cohort analyses

To examine whether SEP across life had cumulative associations with outcomes, analyses in birth cohorts were conducted before and after mutual adjustment for childhood SEP (father’s social class), own education, and own social class at 42/46 years; mutual independence of association after adjustment provides evidence for a cumulative association. In further analyses we used the combined score for father’s social class and own education in the regression models: a larger effect size for the combined score versus either in isolation also suggests a cumulative association.

### Inequalities across the outcome distribution

To examine if the magnitude of inequalities differed across the BP distributions, conditional quantile regression was used.^51^ Quantile regression facilitates estimation of inequalities at a given quantile of the distribution. Using the same ridit scores as described above, estimates of the SII were obtained and plotted at the 5^th^, 10^th^, 25^th^, 50^th^ (median), 75^th^, 90^th^, and 95^th^ quantiles. To facilitate interpretation we show the quantile corresponding to the BP threshold for initiating treatment (140mmHg and 90mmHg for SBP and DBP respectively).

### Additional and sensitivity analyses

To examine how medication use affected the direction and/or magnitude of inequalities in BP, associations between education and BP lowering medication use were analysed using linear probability models, before and after adjustment for SBP (to indicate treatment need). Main analyses were repeated without adjustment for use of BP lowering medication in order to estimate inequalities in observed rather than underlying BP. To examine the robustness of associations of early life SEP with BP—to a different parental respondent and different dimension of disadvantage—maternal educational status was used instead of father’s social class. This was measured via a binary indicator of whether the mother had left formal education at the mandatory leaving age (14 years old from 1918, 15 from 1944, and 16 from 1972). To examine whether differences in missing outcome data may have biased cross birth-cohort study estimates, analyses were repeated using multiple imputation to additionally impute BP outcomes in the eligible target population (i.e., those alive and who had not emigrated). Finally, gender-specific analyses were conducted to examine if any change over time in BP inequalities differed by gender. Likewise, age-specific analyses (25-54, 55+) using the HSE datasets was conducted to examine if age modified the magnitude of inequalities.

### Patient and public involvement

This is a secondary data analysis study which uses data collected from cohort and cross-sectional studies made available for academic research. Patients and the public were not directly involved. Dissemination to study participants and patient organizations is not applicable.

## Results

Comparing 2016 with previous years, both cohort and HSE datasets indicated increased higher educational attainment, more prevalent use of BP lowering medication, and lower mean SBP and DBP (Table 1; differences in SBP in cohort data were more modest than those observed in HSE). Cohort data indicated that the percentage of participants with fathers of manual social class decreased across time (Table 1). The prevalence of outcome missingness in the cohorts, attributable to survey loss to follow-up, migration, and premature mortality from birth to 42-46 years, was higher in each subsequent cohort (40.6% in 1946c, 50.1% in 1958c, and 57.7% in 1970c).

**Table 1.**
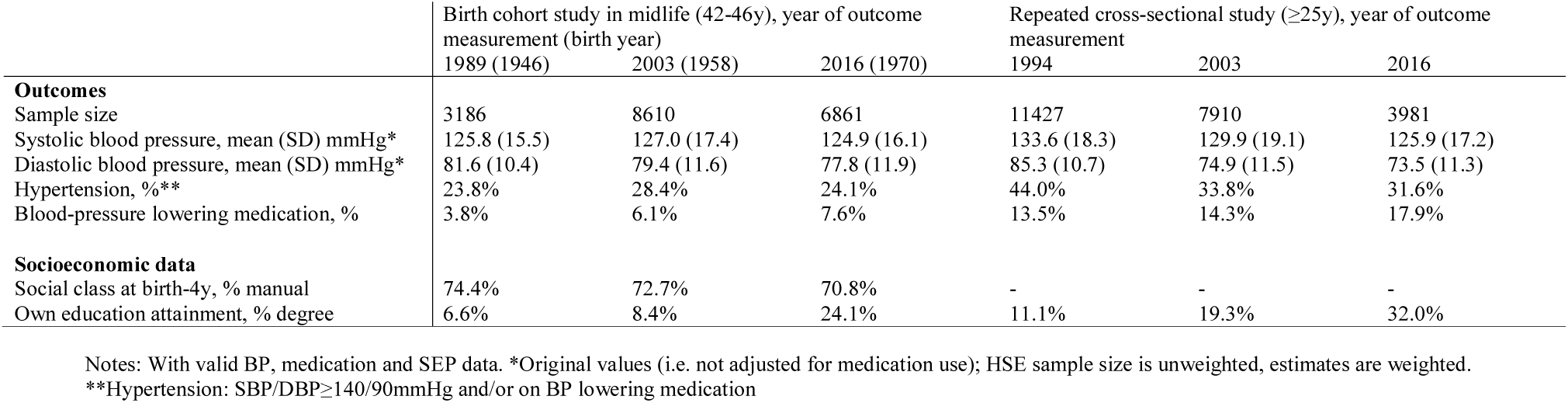
Participant characteristics: data from 3 British birth cohort studies and 21 repeated cross-sectional English studies.

|  | Birth cohort study in midlife (42–46y), year of outcome measurement (birth year) |  |  | Repeated cross-sectional study (≥25y), year of outcome measurement |  |  |
| --- | --- | --- | --- | --- | --- | --- |
|  | 1989 (1946) | 2003 (1958) | 2016 (1970) | 1994 | 2003 | 2016 |
| **Outcomes** |  |  |  |  |  |  |
| Sample size | 3186 | 8610 | 6861 | 11427 | 7910 | 3981 |
| Systolic blood pressure, mean (SD) mmHg* | 125.8 (15.5) | 127.0 (17.4) | 124.9 (16.1) | 133.6 (18.3) | 129.9 (19.1) | 125.9 (17.2) |
| Diastolic blood pressure, mean (SD) mmHg* | 81.6 (10.4) | 79.4 (11.6) | 77.8 (11.9) | 85.3 (10.7) | 74.9 (11.5) | 73.5 (11.3) |
| Hypertension, %** | 23.8% | 28.4% | 24.1% | 44.0% | 33.8% | 31.6% |
| Blood-pressure lowering medication, % | 3.8% | 6.1% | 7.6% | 13.5% | 14.3% | 17.9% |
| **Socioeconomic data** |  |  |  |  |  |  |
| Social class at birth-4y, % manual | 74.4% | 72.7% | 70.8% | - | - | - |
| Own education attainment, % degree | 6.6% | 8.4% | 24.1% | 11.1% | 19.3% | 32.0% |

Notes: With valid BP, medication and SEP data. \*Original values (i.e. not adjusted for medication use); HSE sample size is unweighted, estimates are weighted.
\*\*Hypertension: SBP/DBP≥140/90mmHg and/or on BP lowering medication

### Trends across time in socioeconomic inequalities in BP

In both cohort and HSE datasets, lower education was associated with higher SBP (Figure 1). In HSE, the difference (SII) in SBP based on data pooled across survey years was 4.1mmHg (95% CI: 3.7, 4.5). The magnitude of the social gradient in BP changed little across time. For example, in HSE the SII in SBP was 3.0mmHg (95% CI: 1.8, 4.2) in 1994 and 4.3mmHg (2.3, 6.3) in 2016. Estimates for 1946c were somewhat smaller than 1958c and 1970c, yet 95% CIs overlapped; Figure 1. Disadvantaged early life social class was also associated with higher SBP in each cohort, with similar patterns of association as with education (Figure 1). Findings of persistent inequalities across time in SBP were similar for DBP, and for survey-defined hypertension (Supplementary Figure 1).

**Figure 1.**
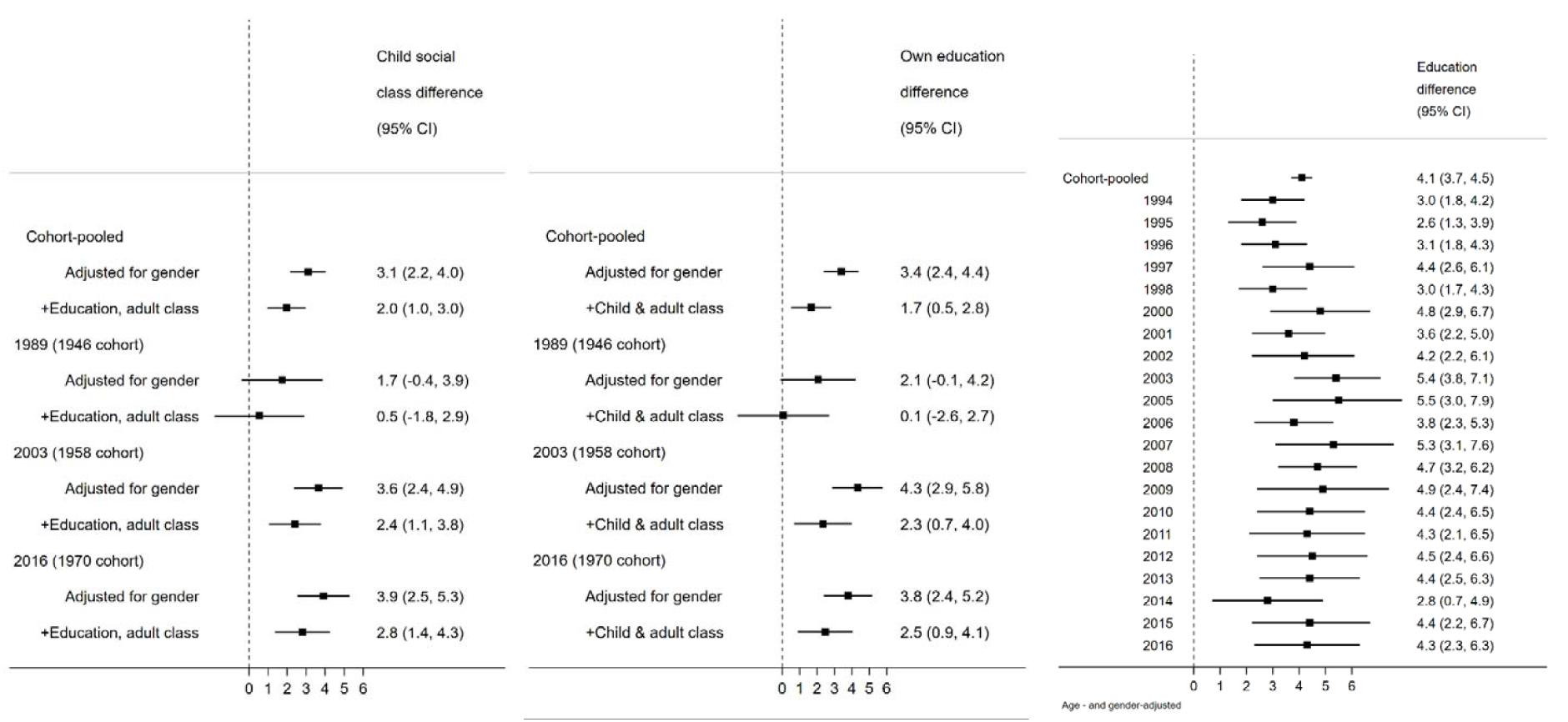
Socioeconomic position across life and mean difference in systolic blood pressure (mmHg) in midlife (42-46 years, from birth cohort data, left panels) and across adulthood (≥25 years, from repeated cross-sectional data, far right panel). Note: estimates are the Slope Index of Inequality (absolute difference in SBP between the lowest and highest socioeconomic position).

### The life course nature of inequalities in BP

More disadvantaged childhood social class and lower own educational attainment were associated with higher SBP, before and after adjustment for each SEP indicator and own adult social class (Figure 1). These adjusted associations were weaker in 1946c than subsequent cohorts, yet 95% CIs overlapped, and as expected, were most precisely estimated in cohort-pooled models (Figure 1, top row). Associations between the composite score (early life social class and own education) and SBP yielded higher effect estimates than either in isolation: in a cohort-pooled model, the difference in SBP was 4.9mmHg (3.7, 6.1) using the composite score yet 3.4mmHg (2.4, 4.4) when considering education alone, and 3.1mmHg (2.2, 4.0) when considering early life social class alone (Figure 1). Findings were similar for DBP and survey-defined hypertension (Supplementary Figure 1).

### Inequalities across the distribution of BP

In both HSE and birth cohorts, quantile regression analyses revealed SEP differences in SBP and DBP across the BP distributions—below and above hypertension thresholds. However, the observed mean differences in SBP and DBP were likely driven by the larger effect sizes at the upper tail of the distribution (Figure 2). For example, in a cohort-pooled model the mean difference in SBP by educational attainment was 1.3mmHg (0.1, 2.6) at the 10^th^ quantile, 2.8mmHg (1.7, 3.9) at the median, and 5.6mmHg (3.4, 7.9) at the 90^th^ quantile (Figure 2). Estimates from pooled HSE analyses were 2.1mmHg (1.6, 2.6), 3.9mmHg (3.5, 4.3) and 5.6mmHg (4.9, 6.4), respectively. We found similar results albeit with lower precision when conducted in each birth cohort and HSE dataset separately; data available upon request.

**Figure 2.**
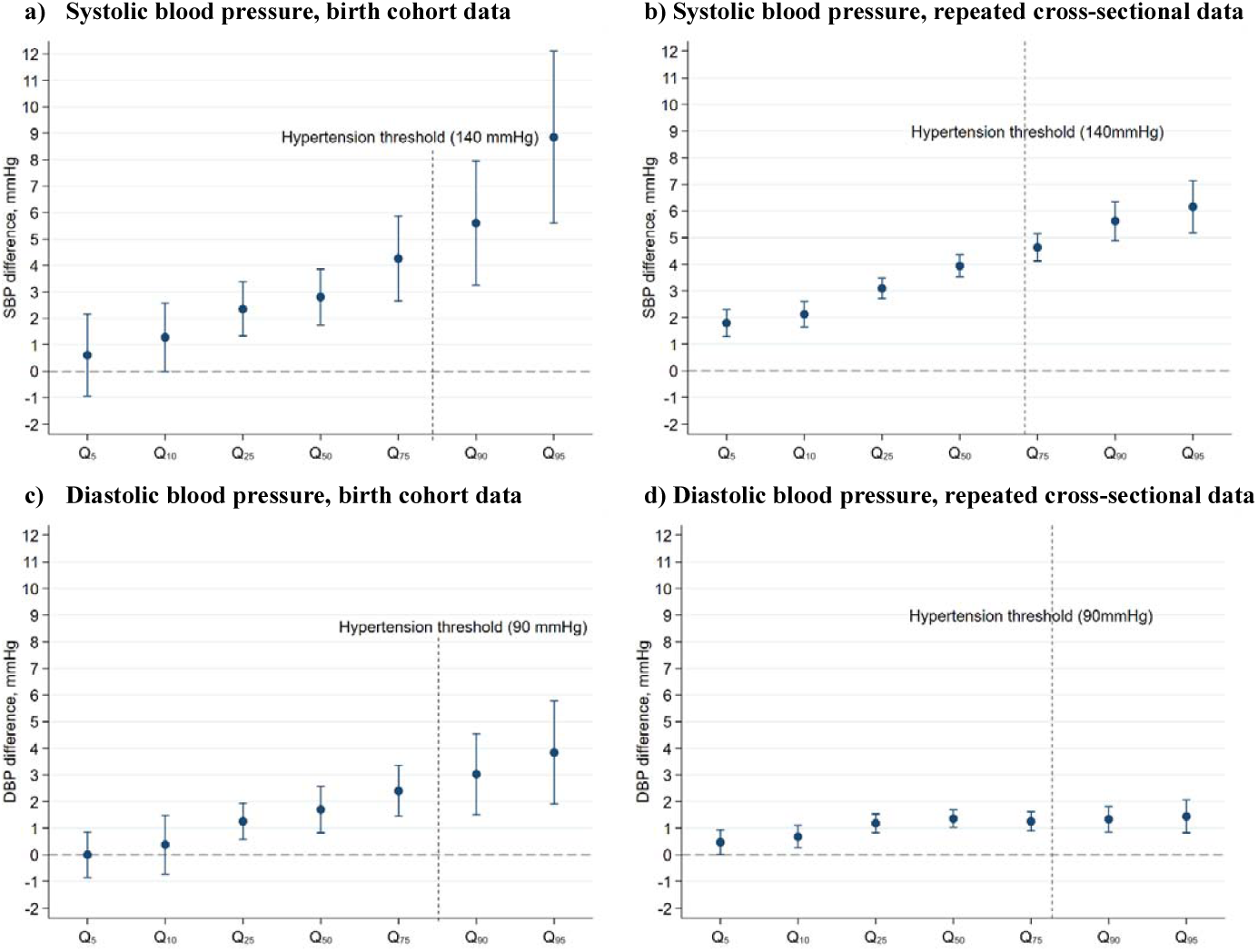
Estimated differences in systolic and diastolic blood pressure in the lowest verses highest education attainment (slope index of inequality): quantile regression estimates at different quantiles of the outcome distribution (95% CI). Coefficients are interpreted analogously to linear regression: for example, Q_50_ shows the median difference in SBP comparing the lowest with highest education attainment. Q5 estimate for DBP in birth cohorts: the model did not converge due to insufficient outcome variance at that quantile.

### Additional and sensitivity analyses

Lower education was associated with increased likelihood of using BP lowering medication (before and after adjustment for measured SBP)—the education related (SII) difference in treatment use after adjustment for SBP was 3.9 (2.4, 5.2) percentage points in a cohort-pooled model and 4.6 (3.9, 5.3) in HSE (Supplementary Figure 2). Main findings were similar when not adjusting BP levels for use of BP lowering medication, yet the magnitude of SEP differences in SBP—as expressed by the SII—was lower in magnitude in each data source (Supplementary Figure 3). Findings were also similar when maternal education was used as an indicator of early life SEP instead of father’s social class—in a birth cohort-pooled model, the SEP difference in SBP according to maternal education was 2.6mmHg (1.7, 3.6), and 6.1mmHg (4.9, 7.3) in a score combining maternal and own education. Finally, findings were also similar when imputing cohort SBP data (data available upon request), and when conducted separately in men and women (Supplementary Figure 4; in HSE, the magnitude of SEP differences in SBP were larger in females, but the persistence of associations across survey years was found in both genders). Exploratory analyses in the pooled and year-specific HSE datasets by age group (25-54, 55+) suggested that age did not modify the magnitude of inequalities (Supplementary Figure 5).

## Discussion

### Main findings

Using data from three birth cohorts and 21 repeated cross-sectional surveys—with data spanning 1989 to 2016—we found persisting socioeconomic inequalities on the absolute scale in adult SBP, DBP, and survey-defined hypertension. We also found, using birth cohorts, evidence for persisting cumulative associations of more disadvantaged SEP across life on these outcomes. Finally, our use of quantile regression found that average differences in SBP and DBP were driven by SEP differences in the upper tail of the BP distributions—i.e., higher inequalities amongst those at highest risk.

### Comparison with previous findings

Our findings of persistent inequalities in BP extend prior analyses of area-based SEP differences in high BP using HSE (1994-2008),^12^ and studies which identified mean differences in BP using 1946c or 1958c.^23-26^ Our effect estimates for 1946c were smaller than those previously reported, a finding which we found attributable to our use of weights to account for the stratified sampling design in this cohort.^33^ While a previous study using 1946c concluded that child SEP may be particularly important for adult SBP (among men),^24^ it used social class only (not education), and a different statistical approach. Our co-ordinated analyses found consistent evidence—across 1958c and 1970c and among both genders—for cumulative associations of early life social class and adult education on BP outcomes.

A recent meta-analysis suggested that SEP and high BP associations were mainly evident for associations with educational attainment;^13^ our findings, using prospectively assessed SEP data across life, suggests that sizable inequalities exist net of education and adult social class. The magnitude of inequalities in BP outcomes are therefore underestimated when solely examined using education as a marker for SEP; this may have implications for future studies which seek to either monitor or reduce inequalities in BP outcomes.

### Explanation of findings

Effective policies to reduce inequalities improve health across all groups but with higher levels of improvement amongst the disadvantaged groups. The apparent persistence of inequalities observed in our study suggests that the population-level declines in average levels of BP from 1989 to 2016 (e.g., 7.7mmHg and 11.8mmHg for SBP and DBP respectively based on HSE data; Table 1) did not substantially differ by SEP. Multiple policies have been enacted across this period which could have feasibly affected such inequalities. These include the attempts to standardise and improve the management of chronic diseases such as BP screening in 2009 (the NHS Cardiovascular Health Check programme in England for those aged 40-74 years), and the introduction of financial incentives for UK general practitioners to regularly monitor BP in hypertensive patients in 2004.

Regardless of the impact of each specific policy,^52 53^ or SEP differences in the management of hypertension,^54^ there are multiple reasons why increases in detection and treatment of high BP are unlikely sufficient to reduce SEP inequalities in BP: 1) treatment is not applicable to those below the BP thresholds for initiating anti-hypertensive medication, in which SEP differences in BP exist (Figure 1); 2) amongst adults with hypertension, those of more disadvantaged SEP are likely to have higher BP levels, such that treatment may not fully normalise BP levels across SEP groups; and 3) across the studied period, inequalities in other factors that may mediate SEP and BP associations, such as higher BMI^12 19^ and unhealthy diets, or are indications for BP lowering treatment, such as diabetes, have likely stubbornly persisted over the last two decades, and these are expected to lead to inequalities in BP.^7^ Consistent with this reasoning, we found that SEP differences in BP were still present, albeit slightly weaker, when treatment for high BP was not accounted for. Additional pathways which may persistently link SEP and BP include psychosocial or physiological processes (e.g., chronic inflammation and stress).^55^ Both early life and adult SEP appear to have cumulative independent associations with such determinants (e.g., BMI^19^) which likely results in cumulative associations with BP in midlife. Future reductions in BP inequalities are likely to be achieved by addressing these wider determinants beyond detection and treatment which influence change in the upper tail of the BP distribution without having a major impact on average levels; these in turn are likely to require structural changes to address, such as those incentivising lower salt and calorie contents of foods.^56^

It is also possible that the persisting BP inequalities found in our study reflect non-causal effects of SEP. For example, those of lower SEP may be increasingly selected on the basis of ill health or factors such as cognition which predict worse subsequent health.^57 58^Such changes may counter-act other favourable changes which would otherwise reduce the magnitude of the observed SEP differences in BP. However, to the authors’ knowledge there is little empirical evidence that selection differs markedly across time—SEP differences in childhood cognition for example appear to have been similar in 1958c and 1970c.^59^

### Strengths and limitations

Strengths of this study include the use of multiple comparable nationally representative datasets with complementary advantages. Our co-ordinated analyses of these enabled investigation of long-run trends in SEP inequalities in BP, the incorporation of data on BP lowering medication, and the use of multiple indicators of SEP collected in both early and adult life. The use of a single analytical framework across the 1946c and 1958c, and additionally including the 1970c, improved the available statistical power and generalisability of our investigation of the life course nature of BP inequalities. Finally, our use of quantile regression enabled us to move beyond a focus on average levels or binary outcomes to identify that such inequalities were driven by differences at the upper tail of the BP distributions.

While we aimed to optimise the comparability of exposure and outcome data, and minimise the potential impact of missing data (via multiple imputation or weighting), we were unable to fully exclude the potential that findings, and thus inferences regarding change in the SEP and BP association across time, are biased due to unobserved predictors of missing data, or unobserved methodological differences between the studies. A further limitation is that, as in other large-scale datasets such as UK Biobank, the study samples were largely White; we lacked power to investigate whether inequalities and their change across time are modified by ethnicity. Finally, while multiple indicators of SEP were utilised, we were unable to investigate unmeasured aspects of socioeconomic circumstances which may be particularly important—such as income-related food insecurity or wealth.

## Conclusions

From 1989 to 2016, socioeconomic inequalities in BP appear to have persisted in absolute terms, with additive associations of early life social class and own education. Indeed, studies examining inequalities in BP solely using education as a marker for SEP are likely to underestimate lifetime inequalities in BP outcomes. This persistence likely reflects the net effects of secular increases in socially patterned drivers of high BP such as higher BMI and diabetes coupled with improved disease management. Assuming consistent causal relations between SEP and BP, our findings suggest that prior strategies of reducing socioeconomic inequalities in these outcomes have been insufficiently effective. Alternative strategies, targeting the wider structural determinants of high BP are likely required.

## Data Availability

1946c data are available from: https://www.nshd.mrc.ac.uk/data/data-sharing/ 1946c, 1958c, 1970c and HSE data are available from the UK Data Archive: https://www.data-archive.ac.uk.

https://www.nshd.mrc.ac.uk/data/data-sharing/

## Acknowledgments

DB is supported by the Economic and Social Research Council (grant number ES/M001660/1) and The Academy of Medical Sciences / Wellcome Trust (“Springboard Health of the Public in 2040” award: HOP001/1025). The HSE is funded by NHS Digital; SS is funded to conduct the annual HSE. RH is Director of CLOSER which is funded by the Economic and Social Research Council (award reference: ES/K000357/1). The funders had no role in study design, data collection and analysis, decision to publish, or preparation of the manuscript. The authors thank the interviewers and nurses, and the participants in the birth cohorts and the Health Survey for England series.

## Contributors

DB and SS conceptualized the study. DB wrote the first draft. DB and SS analysed and interpreted data. DB and SS ACF accept full responsibility for the work and the conduct of the study, had access to the data, and controlled the decision to publish. All authors have approved the final draft of the manuscript.

## Ethical approval

All studies used in this manuscript have received relevant ethical approval; additional ethical approval was not required for the secondary data analysis conducted.

## Data sharing

1946c data are available from: https://www.nshd.mrc.ac.uk/data/data-sharing/ 1946c, 1958c, 1970c and HSE data are available from the UK Data Archive: https://www.data-archive.ac.uk. The lead author (DB) affirms that the manuscript is an honest, accurate, and transparent account of the study being reported; that no important aspects of the study have been omitted; and that any discrepancies from the study as originally planned (and, if relevant, registered) have been explained.

